# “A System Dynamics Model for Long-term Aspect of Caries Preventive Interventions”

**DOI:** 10.1101/2023.04.06.23288258

**Authors:** Tin Htet Oo, Sukanya Tianviwat, Songchai Thitasomakul, Phongpat Sontamino

## Abstract

**Objective:** The purpose of the study is to appraise the long-term effects of caries preventive interventions of the Ministry of Public Health (MOPH), Thailand: supervised toothbrushing (STB), dental sealant, and combined STB+sealant that are implemented for 6- to 15-year-olds by comparing with the base case using the System Dynamics Model (SDM).

**Method:** The System Dynamics Model (SDM) was analyzed to compare the interventions, supervised toothbrushing (STB), sealant, and combined STB+sealant as intervention scenarios with the base case scenario. The effectiveness data of interventions set in the model were retrieved from meta-analyses and experts’ opinions.

**Results:** The model established that the no caries population increased by 36.2%, 25.5%, and 18.6% while the caries population decreased by 8.1%, 5.6%, and 3.3% under combined STB+sealant, sealant, and supervised toothbrushing (STB) respectively compared to the base case at the age of 15 years old. From the above 20 years old to the elderly age, both individual and combined interventions have no favorable effect on reducing caries. On the other hand, the no caries population under the interventions is not significantly different from the base case during the elderly age.

**Conclusion:** Combined STB+sealant is the most effective intervention among the interventions that are administered during the age of 6-15 years. The interventions given during these ages have effectiveness in caries reduction by about 15 years from when they are started. In addition, System Dynamics Model (SDM) would assist in comparing the interventions or policies as scenarios to indicate the most opportune one for the population.

## Introduction

Dental caries is still regarded as a burden of oral disease not only in Thailand but also around the world even though it is preventable [1]. Studies have shown a reduction in the prevalence of dental caries over decades in developed countries [2, 3]. However, in several developing countries, there has been an increasing trend in caries prevalence reported by some studies [3, 4]. The high prevalence of dental caries in developing countries is due to poor oral hygiene habits, low awareness of caries, growth in sugary consumption, insufficient access to preventive programs for dental health, and inadequate exposure to fluoride [3]. According to the 8^th^ National Oral Health Survey (NOHS) in Thailand in 2017, the prevalence of dental caries was still high at 52% in 12-year-olds, 62.7% in 15-year-olds, 91.8% in 35- to 44-year-olds, 98.5% in 60- to 74-year-olds, and 99.5% in 80- to 85-year-olds [5]. People having dental caries could suffer from pain, disturbance during eating, aesthetics, and functional problems [6]. Moreover, tooth loss due to caries was the most common consequence of caries [7].

Due to the burden of caries, the role of prevention and promotion of oral health is vital. Therefore, various preventive programs such as oral health education, toothbrushing, fluoride varnish, application of dental sealant, and so on have been designated for dental caries in both individuals and communities [8-11]. In Thailand, the Ministry of Public Health (MOPH) proposed preventive oral health programs for respective age groups including toothbrushing, fluoride varnish, dental sealant, oral health examination, and so on [12]. However, a long-term assessment of preventive interventions for conducting better decision-making and an alternative program is needed. Simulation models would help policymakers in making decisions for planning long-term processes. [13]. They have been widely used in health care and a study also reported models that were reliable to be used in dental caries interventions such as the Markov Model, System Dynamics Model (SDM), Microsimulation Model, and Decision-tree Model [14].

In this study, SDM was explored for caries preventive interventions intended by the Ministry of Public Health (MOPH) such as supervised toothbrushing (STB), sealant, and their combination. The interventions were primary preventive interventions for caries since they focused on preventing the occurrence of diseases in a healthy state before the process begins [15]. The System Dynamics Model (SDM) is a computer simulation to help the stakeholders in deciding the most admissible policy or intervention for the community [16]. It can be used as a cohort-based model and shows the interaction between the entities of the system by modeling the rate of change of the system [17, 18]. It also allows seeing feedback loops that emerge from the interaction [17, 18]. SDM should be considered as a cohort-based simulation model for the long-term aspect of the outcomes not only under individual preventive intervention but also the combination of the interventions.

Therefore, the objective of the study is to estimate the effectiveness of providing caries preventive interventions for 6- to 15-year-olds by the Ministry of Public Health (MOPH) namely supervised toothbrushing (STB), sealant, and combined STB+sealant, compared with the base case as a long-term aspect by conducting the System Dynamics Model (SDM).

## Methodology

The modeling in this study focused on the permanent dentition age. The System Dynamics Model (SDM) involves two parts: qualitative and quantitative. The qualitative part showed the causal relationship of caries related and identifies the feedback loops among them by a causal loop diagram (CLD). The quantitative part translated the causal loop diagram (CLD) for evaluating the conditions quantitatively represented as stocks and flows diagram. Group model building (GMB) sessions including the researchers and experts in related fields were held three times to determine the final model. The experts are dentists with valuable experience and knowledge in dental public health and clinical fields. The activities such as identifying the time horizon, identifying the key variables, and their behaviors, formulating the causal loop diagram (CLD), and developing the stocks and flows diagram were involved in group model building (GMB) sessions. The setting interventions in the model were implemented interventions by the Ministry of Public Health (MOPH). Since the study was based on the effectiveness of the interventions, systematic reviews and meta-analyses of interventions were conducted first by the researchers for the effectiveness data to be used in the model. The Vensim DSS version 6.4 software was used for running the model. The model simulated the Thai population born in 2021 to 75 years old (average life span) to estimate the outcomes in age intervals under intervention scenarios. The total population was 678,243. The results are presented at the age of 15 years old when the interventions were discontinued and at 60 years old to see the long-term effect on the elderly.

### Systematic review and meta-analysis

Systematic reviews and meta-analyses were conducted by researchers to identify the effect of supervised toothbrushing (STB) and sealant (6-15 years old) for dental caries by following the guidelines of the systematic review of interventions in Cochrane Handbook version 5.1.0. [19]. The included studies were identified according to PICO (Participants, Intervention, Comparison, and Outcome). For the STB, the studies were searched from Cochrane, PubMed, Web of Science, and SCOPUS databases. The protocol was registered at Prospero (CRD42022376887). For sealant, the studies were retrieved from the previous recent review [20]. Meta-analyses with the random-effect model for both interventions were performed by researchers in RevMan 5.3 software. The estimated effect size in the meta-analysis was risk ratio (RR) and effective rates of interventions were calculated by the formula ([1-RR]*100).

### Model structure

The causal loop diagram (CLD) presents the causal relationship of no caries, caries, related events of caries, and feedback loops among them, (Fig 1). A positive sign (+) in the diagram means that a variable adds to another one and a negative sign (-) that it subtracts from the other [21]. One balancing loop and two reinforcing loops were found in the diagram. A balancing loop means that a positive change in a variable lead to a pushback in the opposite direction and if the direction of variables keeps in the same direction, it is a reinforcing loop [21]. A balancing loop in the diagram shows that an increase in the no caries leads to an increase in caries and missing teeth; on the other hand, an increase in the missing teeth leads to a decrease in the no caries. When the intervention is given, it would reduce developing caries, and missing teeth, and on one side, the no caries would increase. Reinforcing loops show that an increase in caries tends to increase in the treated cases due to caries (restoration and endodontic) and recurrent caries from the treated cases. When the intervention is set, it would decrease developing caries, treated cases, and recurrent caries from the treated cases.

**Fig 1.**
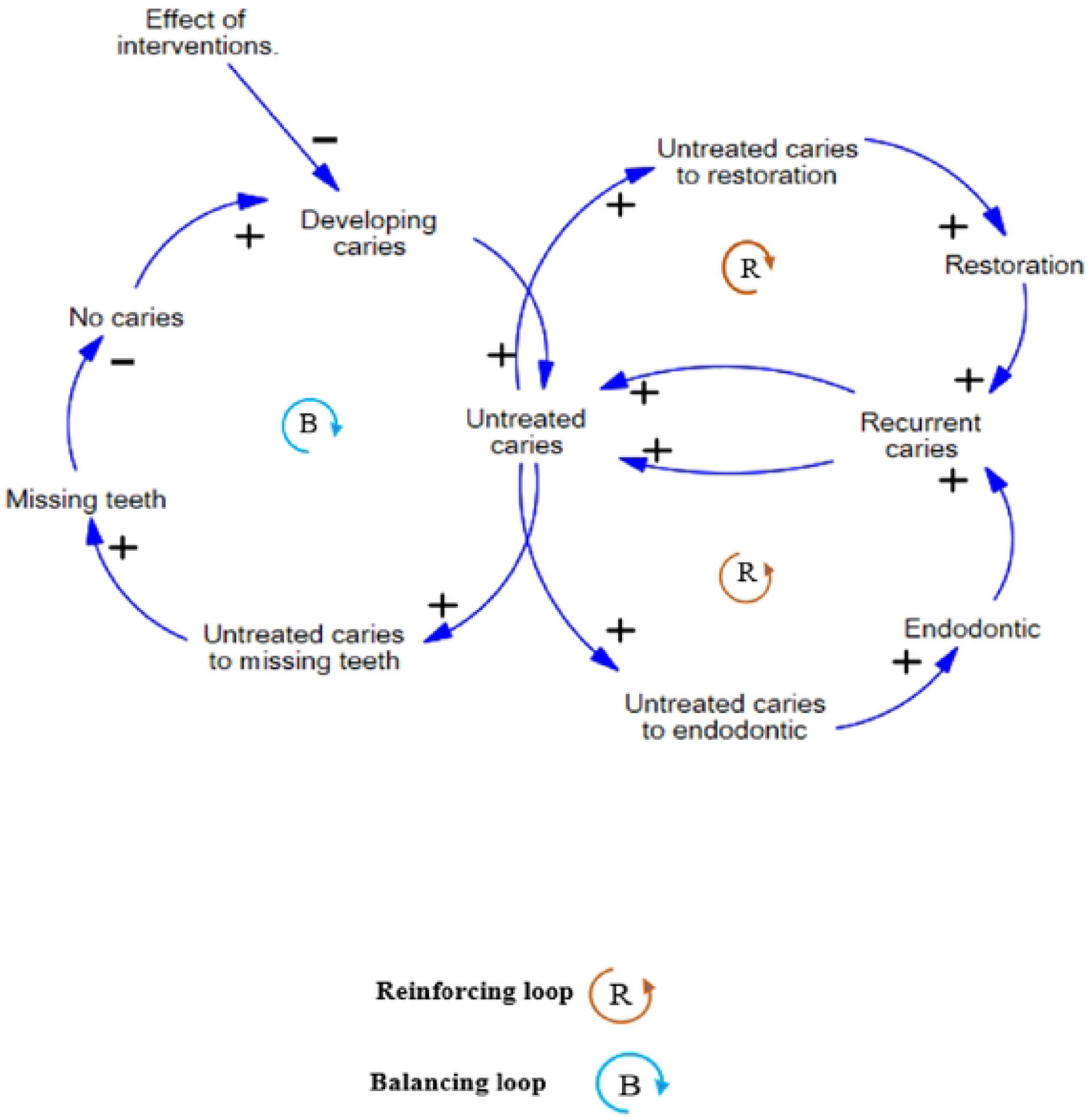
Causal loop diagram (CLD)

CLD was translated into the stocks and flows diagram, Fig 2, to evaluate the outcomes quantitatively. The square blocks in the diagram are denoted as stocks and the arrow symbols represent the flows. The stocks are defined as states or levels of variables moving through the system [13]. The flows control the movement of stocks or determine the changes in the stocks by in and out valves of the stocks [13]. The quantitative outcomes are attributed to the stocks in the model. The fractions affect the rate of flows, for example, the caries development fraction influences the rate of developing caries, and the fraction of restorative treatment affects the rate of untreated caries to restoration. It is assumed that when the intervention is set, the interventions could reduce the caries development fraction tending to slow the rate of developing caries as shown in Fig 2.

**Fig 2.**
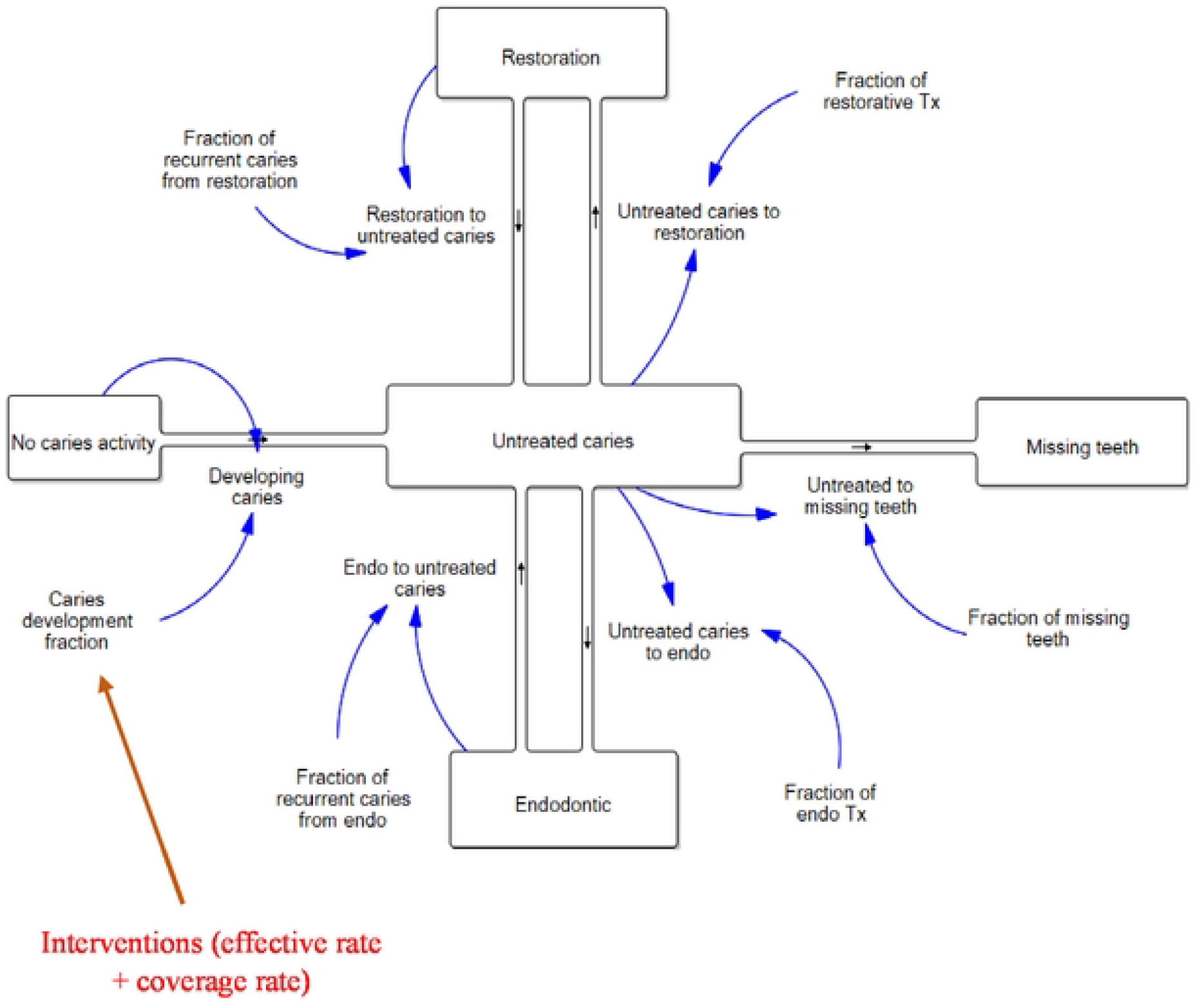
Stocks and Flows diagram.

### Outcomes of the model

The stocks in the model defined the main outcomes: no caries and caries and also consequences of caries as follows –

- Population with no caries
- Population with untreated caries
- Population with restorative treatment
- Population with endodontic treatment
- Population with missing teeth

### Source of data

The effective rates of the interventions to be used in the model were obtained from systematic reviews and meta-analyses as mentioned above. The other required parameters in the model were obtained from Thailand’s data: the 8^th^ National Oral Health Survey (NOHS), the Health Data Center (HDC), the National Statistical Office, and literature, and experts’ opinions. When data were not available, experts’ opinions were consulted. Data used in the model were validated by experts from the group model building (GMB) session.

### Scenario analysis

The interventions were analyzed as intervention scenarios comparing the base case scenario as follows. It is assumed that the interventions were provided at 6-15 years old and did not continue after 15 years old. The effective rate remained unchanged after that age.

### Base case

The base case scenario refers to the situation in which no intervention was given. All model parameters in the base case remained unchanged over the simulation run. It served as a reference point for comparing intervention scenarios.

### Supervised Toothbrushing (STB)

It aims to prevent the development of dental caries using the correct toothbrushing technique with fluoride toothpaste and appropriate toothbrush design twice daily under the supervision of dental health professionals or trained teachers or parents [9]. The coverage rate of the intervention at 6-15 years old in Thailand was 95% [22]. The effective rate for this age group was retrieved as 10% from the meta-analysis. The adjusted effective rate with the coverage rate (9.5%) was set in the model intended to reduce the caries development rate.

### Dental Sealant

It is purposed to prevent dental caries by applying resin-based sealants in deep pits and fissures of permanent teeth by dental professionals [11]. In Thailand, the coverage rate of sealant from 6-15 years old was 27% [23]. According to a meta-analysis, dental sealant has a 71% caries reduction on molar teeth at these ages. Based on the meta-analysis and experts’ opinions using their available existing data, a dental sealant was regarded 58% decreased risk of caries for all teeth. It was adjusted with the coverage rate and the adjusted effective rate, 15.7% was set in the model to decrease the rate of caries development.

### Combined Supervised Toothbrushing and Sealant (STB+Sealant)

The descriptions of STB and sealant are the same as mentioned above. The coverage rate of the combined STB+sealant in Thailand for 6-15 years old was 33% [22]. The effective rates of STB and sealant before adjusting with the coverage rates were 10% and 58% respectively as mentioned above. According to each rate and experts’ opinions based on their existing data, the effective rate of combined intervention was determined as 60%. After adjusting the coverage rate, the adjusted effective rate of 19.8% was set in the model to slow the caries development rate.

### Model validation

Both structure-based validation and behavior-based validation were conducted. Face validity was conducted for the model’s structure as structure-based validation by researchers and experts in the group model building (GMB) sessions. As a structure-based validation, the adequacy of the model boundary, model’s structures, parameters verification, and dimensional consistency of the model’s equations were checked [24]. Known-case validity compared with historical data and sensitivity analysis by changing the plausible parameters were evaluated by researchers as behavior-based validation for the model’s behavior [24].

### Uncertainty analysis

Uncertainty analysis was done as multivariate sensitivity analysis with random uniform distribution in Vensim DSS version 6.4 software to examine how a change in the parameters would influence the outcomes. The parameters were varied by ±10 percent. The mean values of the outcome (population with no caries) with a 95 percent confidence interval under the base case scenario and intervention scenarios are presented in Tables 1 and 2.

**Table 1.** Uncertainty analysis of the population with no caries at the age of 15 years.

| Scenarios | Population with no caries |  |  |
| --- | --- | --- | --- |
|  | Mean | Lower bound<br>(95% CI) | Upper bound<br>(95%CI) |
| Base-case | 112,383 | 97,023 | 128,402 |
| STB | 133,943 | 117,002 | 149,809 |
| Sealant | 141,006 | 125,046 | 157,423 |
| STB + sealant | 153,526 | 137,157 | 169,079 |

**Table 2.** Uncertainty analysis of the population with no caries at the age of 60 years.

| Scenarios | Population with no caries |  |  |
| --- | --- | --- | --- |
|  | Mean | Lower bound<br>(95% CI) | Upper bound<br>(95%CI) |
| Base-case | 141 | 52 | 282 |
| STB | 338 | 142 | 631 |
| Sealant | 532 | 240 | 957 |
| STB + sealant | 857 | 414 | 1,483 |

### Ethical considerations

This study received ethical approval from the Human Research Ethics Committee of the Faculty of Dentistry, Prince of Songkla University, EC6503-011 on 7 March 2022.

## Results

Table 3, the population with no caries under combined STB+sealant is the highest, 153,042 (22.56%) than in other scenarios: dental sealant, 141,097 (20.80%), supervised toothbrushing (STB), 133,306 (19.65%) and base-case, 112,405 (16.57%) at the age of 15 years. It increased by 36.2% under the combined STB+sealant, 25.5% in sealant, and 18.6% in STB compared to the base case. The untreated caries population is the lowest under combined STB+sealant, 257,655 (37.99 %) followed by sealant, 264,507 (38.99%) and STB, 270,870 (39.94%) whereas the base-case is the highest, 280,244 (41.32%). Compared to the base case, it decreased by 8.1% in combined STB+sealant, 5.6% in sealant, and 3.3% in STB. The population with missing teeth and populations with restoration and endodontic treatment are also the highest in base-case and lowest in combined STB+sealant followed by sealant and STB. Compared to the base case, the population with restoration decreased by 6.1%, 4.4%, and 3.8% while the population with endodontic decreased by 4.7%, 3.3%, and 3.0% in combined STB+sealant, sealant, and STB respectively. The missing teeth population decreased by 6.7% in combined STB+sealant, 4.8% in sealant, and 4.3% in STB compared to the base case.

**Table 3.** Simulation results when the age of 15 years.

| State | Scenarios |  |  |  |
| --- | --- | --- | --- | --- |
|  | Base case<br>(a) | STB<br>(b) | Sealant<br>(c) | STB +<br>Sealant<br>(d) |
| No caries | 112405<br>(16.57%) | 133306<br>(19.65%) | 141097<br>(20.80%) | 153042<br>(22.56%) |
| (% change from the base case) | - | +18.6 | +25.5 | +36.2 |
| Untreated caries | 280244<br>(41.32%) | 270870<br>(39.94%) | 264507<br>(38.99%) | 257655<br>(37.99%) |
| (% change from the base case) | - | -3.3 | -5.6 | -8.1 |
| Restoration | 119813<br>(17.67%) | 115259<br>(16.99%) | 114601<br>(16.89%) | 112547<br>(16.59%) |
| (% change from the base case) | - | -3.8 | -4.4 | -6.1 |
| Endodontic | 12822<br>(1.89%) | 12436<br>(1.83%) | 12390<br>(1.83%) | 12222<br>(1.80%) |
| (% change from the base case) | - | -3.0 | -3.3 | -4.7 |
| Missing teeth | 152959<br>(22.55%) | 146372<br>(21.58%) | 145648<br>(21.47%) | 142777<br>(21.05%) |
| (% change from the base case) | - | -4.3 | -4.8 | -6.7 |

STB (b), % change from the base case = $(b - a) / a * 100$
Sealant (c), % change from the base case = $(c - a) / a * 100$
STB+Sealant (d), % change from the base case = $(d - a) / a * 100$

Table 4, at the age of around 60 years, the no caries population is the lowest population among other populations. Even though there is no significant difference between the scenarios, the highest, 808 (0.12%) is in combined STB+sealant, followed by sealant, 496 (0.073%), and supervised toothbrushing (STB), 311 (0.046%) compared to the base case, 127 (0.019%). Whereas the untreated caries population increased by 4.9% in combined STB+sealant, 3.2% in sealant, and 2.1% in STB compared to the base case. For the population with restoration, it increased by 1.6%, 1.1%, and 0.9% while the population with endodontic decreased by 0.66%, 0.43%, and 0.28% under combined STB+sealant, sealant, and STB respectively compared to the base case. The population with missing teeth decreased by 1.2% in combined STB+sealant, 0.79% in sealant, and 056% in STB from the base case.

**Table 4.**
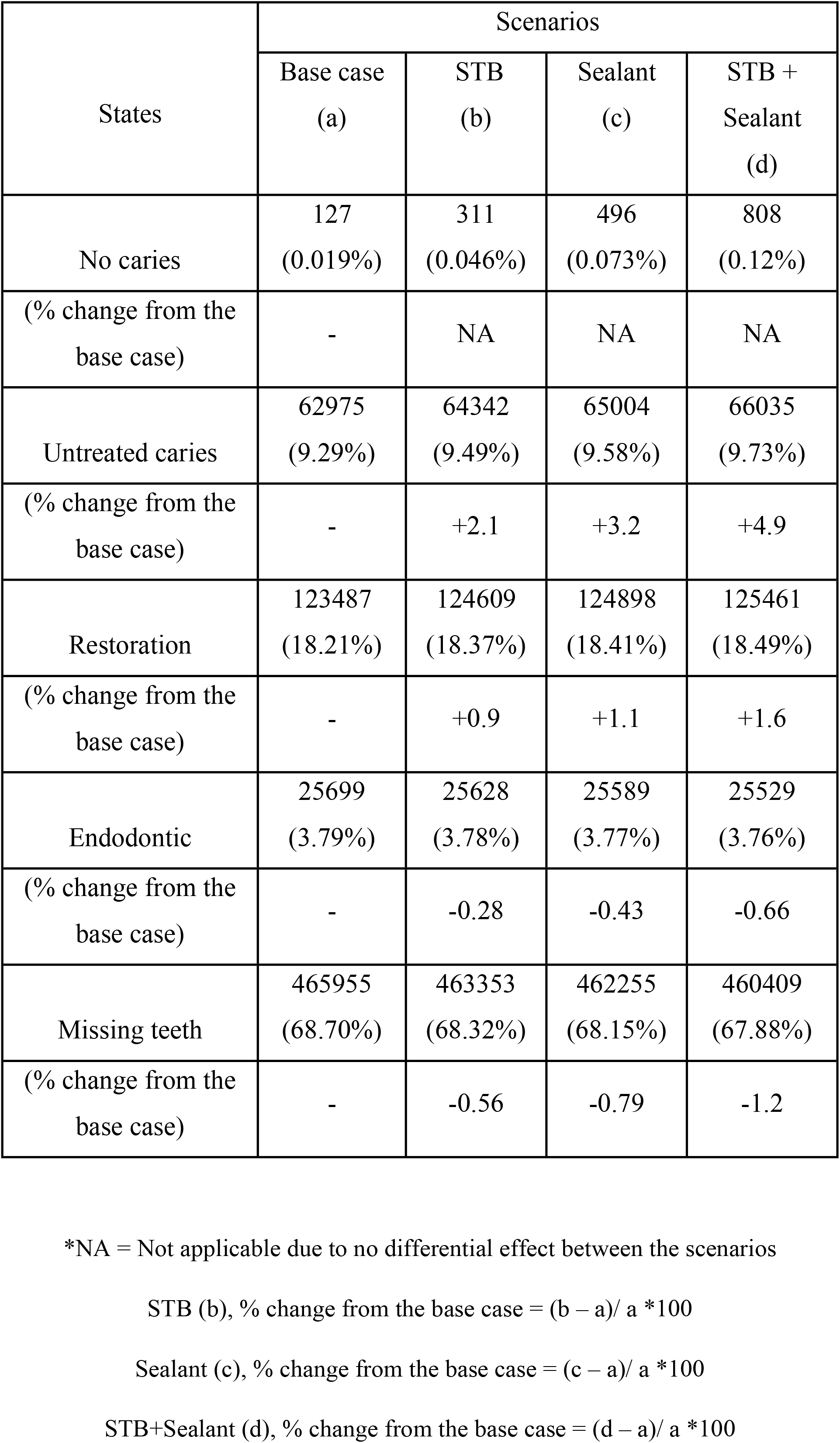
Simulation results when the age of 60 years.

| States | Scenarios |  |  |  |
| --- | --- | --- | --- | --- |
|  | Base case<br>(a) | STB<br>(b) | Sealant<br>(c) | STB +<br>Sealant<br>(d) |
| No caries | 127<br>(0.019%) | 311<br>(0.046%) | 496<br>(0.073%) | 808<br>(0.12%) |
| (% change from the<br>base case) | - | NA | NA | NA |
| Untreated caries | 62975<br>(9.29%) | 64342<br>(9.49%) | 65004<br>(9.58%) | 66035<br>(9.73%) |
| (% change from the<br>base case) | - | +2.1 | +3.2 | +4.9 |
| Restoration | 123487<br>(18.21%) | 124609<br>(18.37%) | 124898<br>(18.41%) | 125461<br>(18.49%) |
| (% change from the<br>base case) | - | +0.9 | +1.1 | +1.6 |
| Endodontic | 25699<br>(3.79%) | 25628<br>(3.78%) | 25589<br>(3.77%) | 25529<br>(3.76%) |
| (% change from the<br>base case) | - | -0.28 | -0.43 | -0.66 |
| Missing teeth | 465955<br>(68.70%) | 463353<br>(68.32%) | 462255<br>(68.15%) | 460409<br>(67.88%) |
| (% change from the<br>base case) | - | -0.56 | -0.79 | -1.2 |

\*NA = Not applicable due to no differential effect between the scenarios
STB (b), % change from the base case = $(b - a) / a * 100$
Sealant (c), % change from the base case = $(c - a) / a * 100$
STB+Sealant (d), % change from the base case = $(d - a) / a * 100$

Figure 3 shows the population with no caries of permanent dentition age, from 6 years old to elderly age. It seems that the no caries population is the highest under combined STB+sealant followed by sealant and supervised toothbrushing (STB) and lowest under base-case. It is still higher under intervention scenarios than the base case until the adult age and the differences can be seen prominently in the graph before the age of 30 years. Nevertheless, it cannot be seen differently between interventions and the base case after the age of 40 years. Besides, it is seen that the population with no caries significantly decreases over time.

**Fig 3.**
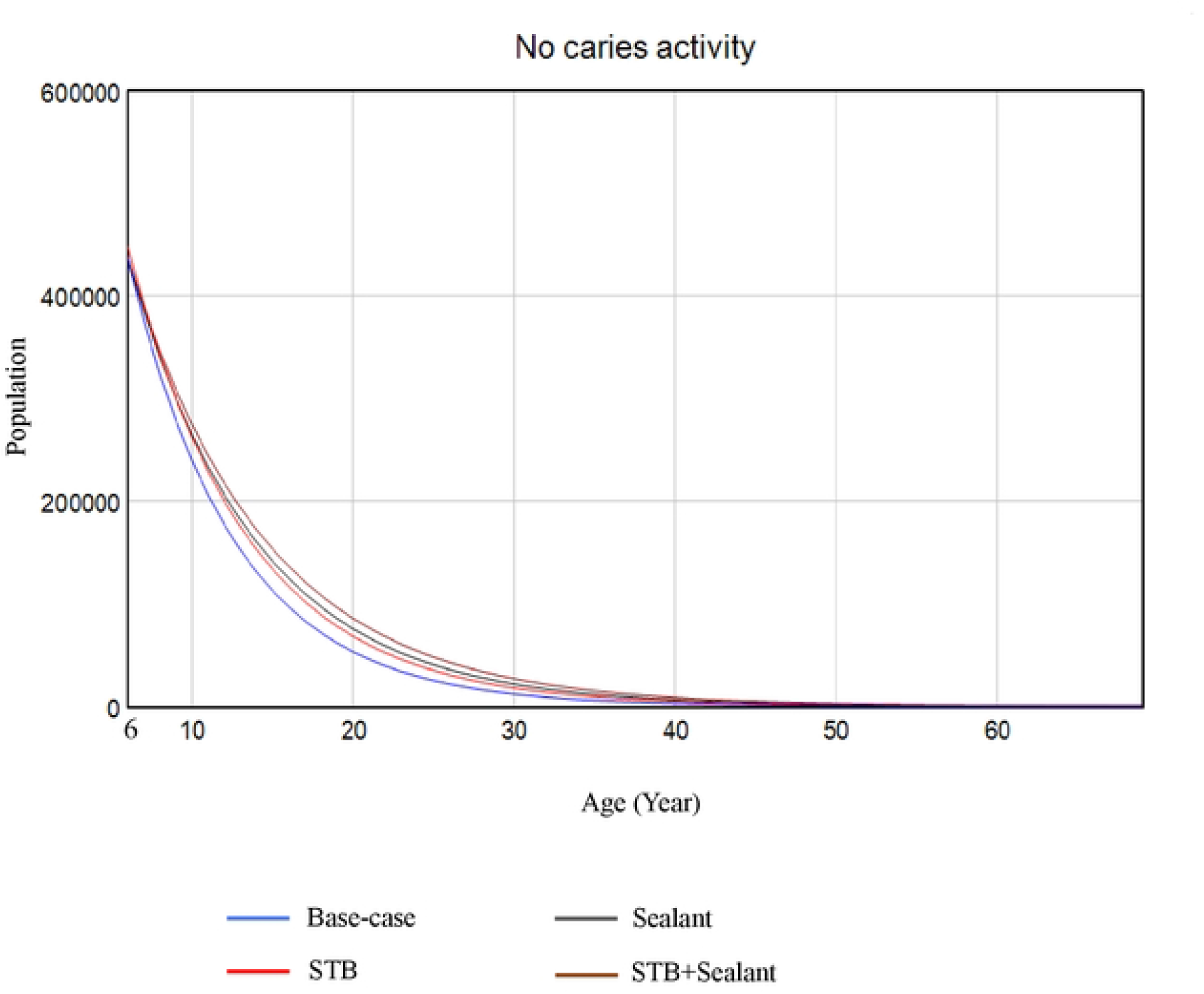
Population with no caries.

Figure 4 presents the untreated caries population from 6 years old to elderly age. It shows that it is the lowest under combined STB+sealant followed by sealant and STB then base-case. It can be seen clearly in the graph until the age of 20 years. However, at ages over 20 years, the caries population under interventions is not lower than the base case. Further, the graph shows that the caries population is slightly decreased over the years.

**Fig 4.**
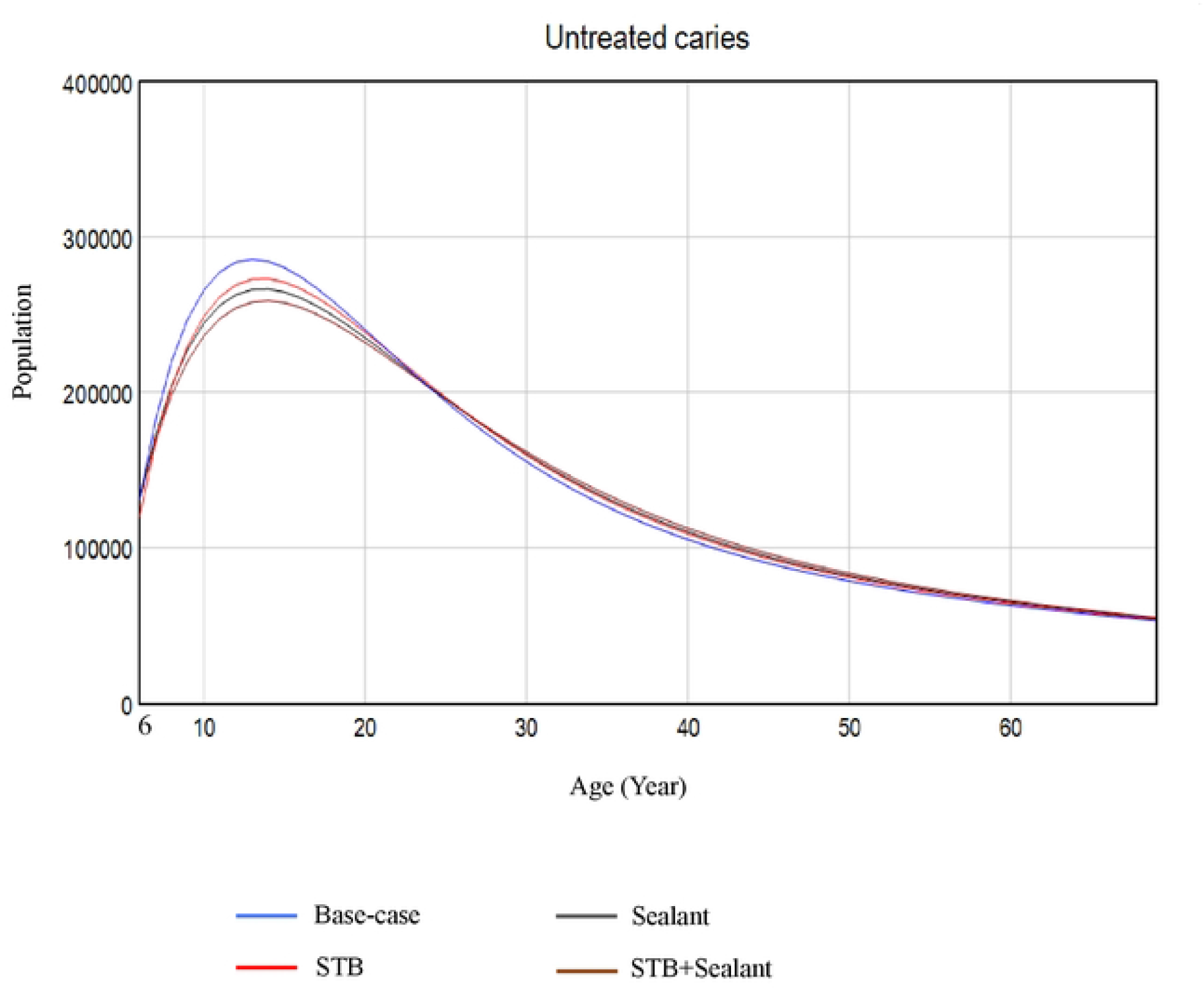
Population with untreated caries.

## Discussion

When establishing the interventions in the model, the effective rates of the interventions were identified and adjusted from the coverage rates in Thailand. Depending on the coverage rate, the effective rate of supervised toothbrushing (STB) was only 9.5%. Although the coverage rate is high, the effective rate was not high, and the caries development rate was not significantly reduced from the base case. While the sealant and combined STB+sealant were quite good in effective rates, the coverage rates were not satisfactory, and their adjusted effective rates were 15.7% and 19.8% respectively. It is considered that the effective rates were not very high due to quite low coverage rates. Therefore, the reduction of caries development rate under these two interventions was also not significantly different from the base case.

Since the effective rate of supervised toothbrushing (STB) is not very satisfactory, this must be emphasized in the intervention. A study indicated that STB has a favorable effect on preventing caries [25]. In this study, the children received an intensive preventive program including the Bass method of brushing and finger flossing monitored by dentists. During the program, daily practicing toothbrushing with fluoride toothpaste in addition to flossing was carried out after lunch on every school day under the supervision of school nurses for one semester (20 weeks). This program lasted about 10 years. Therefore, the type of resource used, and the scheme would be crucial for the intervention, and a comprehensive approach to toothbrushing should be contemplated for more beneficial effects. In the aspect of sealant, applying the resin-based sealant on the pit and fissure permanent teeth has a valuable effect on caries. However, more resources would be needed than for STB, and the availability of resources needs to be of concern.

Based on the simulation, combined STB+sealant is better than each of these assigned separately. Supervised toothbrushing (STB) is proposed not only to prevent the occurrence of dental caries but also to develop oral hygiene habits [9] while the dental sealant is especially focused on molar teeth with a quite good preventing effect on caries [11]. Therefore, the combination of the interventions would be better intervention. It can be seen that at the age of 15 years, under the combined intervention, the no caries population is the highest, and the populations with untreated caries, restoration, endodontic treatment, and missing teeth are the lowest compared with each intervention administered alone, as shown in Table 3. It also seems that the no caries population remains the highest under the combined intervention among the adult and elderly ages.

Even though the no caries population is still higher than in the base case until the elderly age under intervention scenarios, the untreated caries population gradually increased in intervention scenarios over the base case scenario after 20 years old. It is regarded that the interventions that are allocated among the younger ages could no longer reduce the untreated caries population in the adult ages. Therefore, the interventions have a caries reduction effect for only about 15 years from when they are started. It shows that the consequences of caries such as treated cases due to caries and missing teeth under intervention scenarios are not very different from the base case in the elderly ages since the interventions have no positive effect on caries from the adult age.

A study by Splieth et al., (2008) [26] developed the System Dynamics Model (SDM) and reported the combination of fluoride toothpaste, fluoride salt, and fluoride gel was the most effective among fluoride regimes for caries prevention and was also cost-effective. Another study (Hirsch et al., 2012) [27] reported that the SDM estimated the combined early childhood caries (ECC) interventions were the most significant approach for caries reduction and costs. The present study is consistent with these previous studies in that combined interventions are preferred based on SDM simulation. Two studies (Hirsch et al., 2012, Edelstein et al., 2015) projected caries reduction for 10 years by simulating the SDM [27, 28]. It is analogous to this study where the reduction of caries persists for about 15 years. Nevertheless, the present study focused on the long-term simulation of more than the projected 10 years. Furthermore, the previous studies mentioned only the effective rates of the interventions whereas this study concerned not only the effective rates of the interventions conducted by meta-analyses but also the coverage rates in Thailand.

In this study, the data for the cause of missing teeth due to dental caries or periodontal disease was impossible to separate depending on the available data source, Health Data Centre (HDC). Therefore, the data of all causes of missing teeth from HDC were used and it was assumed that the missing teeth may be mostly due to caries among the younger ages, and from both caries and periodontal diseases during the adult and elderly ages [29, 30]. Therefore, if the interventions focus not only on caries but also on the periodontal disease with huge effectiveness, the number of the population with missing teeth could be reduced more than that indicated in this simulation study. In contrast, it would not be different if the focus were to be on caries alone. Moreover, considering each socioeconomic factor related to dental caries as a variable in the model is complicated. It is assumed that developing caries is prevented by reducing the risk factors that could be influenced by interventions. Due to the above limitations, this study may require accomplishing a more distinct quantitative model prediction.

Since it has been allowed that combined STB+sealant is more effective to implement, expanding the coverage rate of the intervention must be considered. This is because the effective rate of the combined intervention is quite low after adjusting the coverage rate although the combined intervention is good in effect. Besides, since the interventions afforded among younger ages have meager effectiveness in adult age and elderly age, furnishing some effective interventions beyond the younger ages should be considered.

The System Dynamics Model (SDM) may be used for long-term assessment in analyzing the effectiveness of interventions when projecting the interventions for improving oral health. Moreover, the SDM may help in comparing the cases as “what if” scenarios to anticipate the best case and poor case of other services or policies. Since the study did not encompass the costs, cost-effectiveness, or cost-saving, these aspects should be appraised in further studies.

## Conclusion

Among the interventions provided by the Ministry of Public Health (MOPH) during the age of 6-15 years, the combined STB+sealant is the best intervention compared with each applied separately. It is supposed that the interventions given between the ages of 6-15 years old can reduce caries for 15 years from when they are started but have no beneficial effect on the population during the elderly age. Moreover, System Dynamics Model (SDM) would help decision-makers anticipate the best interventions or policies by simulating scenarios.

## Data Availability

All relevant data are within the manuscript and its Supporting Information files.

## Supporting information

**S1 Fig 1. Causal loop diagram (CLD).**

**S2 Fig 2. Stocks and Flows diagram.**

**S3 Fig 3. Population with no caries**.

**S4 Fig 4. Population with untreated caries**.

**S5 Table 1. Uncertainty analysis of the population with no caries at the age of 15 years**

**S6 Table 2. Uncertainty analysis of the population with no caries at the age of 60 years**

**S7 Table 3. Simulation results when the age of 15 years**

**S8 Table 4. Simulation results when the age of 60 years**

